# Blood Pressure and Hypertension After Hypertensive Disorders of Pregnancy

**DOI:** 10.1101/2025.01.02.25319922

**Authors:** Rachel Peragallo Urrutia, Matthew Shane Loop, Jasmine D Johnson, Tracy Y Wang, Thomas M Price, Melissa A Daubert

**Affiliations:** University of North Carolina School of Medicine, Department of Obstetrics and Gynecology, Division of General Obstetrics, Gynecology, and Midwifery; Department of Health Outcomes Research and Policy, Harrison College of Pharmacy, Auburn University; Division of Pharmacotherapy and Experimental Therapeutics, UNC Eshelman School of Pharmacy, University of North Carolina at Chapel Hill; Indiana University School of Medicine, Department of Obstetrics and Gynecology, Division of Maternal Fetal Medicine; Duke University, Department of Medicine, Division of Cardiology, Duke Clinical Research Institute; Duke University Department of Obstetrics and Gynecology, Division of Reproductive Endocrinology and Infertility

## Abstract

**Introduction:** Cardiovascular disease (CVD) is the leading cause of mortality for women. Timely diagnosis of hypertension after a hypertensive disorder of pregnancy (HDP) provides an opportunity for CVD prevention. We assessed the association between blood pressure (BP) 15-90 days postpartum and incident hypertension after an HDP.

**Methods:** This was a retrospective cohort study of women with an HDP between January 2014 and December 2017 at two health systems in the southeastern U.S. Cox proportional hazards models assessed the association of postpartum BP and incident hypertension 12 months postpartum. Covariates included type of HDP, gestational age at diagnosis, timing of measurement, comorbidities, and structural determinants of health. We excluded people with preexisting hypertension and without a BP measurement 15-90 days postpartum.

**Results:** Out of 5657 women, only 2514 (44%) met the inclusion criteria as almost 40% (2125) did not have a BP check at 15-90 days postpartum. The hazards of incident hypertension were significantly higher for those with elevated systolic postpartum BP (1.70, 95% CI: 1.37 - 2.12) and more severe HDPs. The estimated cumulative incidence of hypertension among participants with postpartum BP of 110/65 mmHg was 4.7% (CI, 2.0% - 7.4%) and for those with BP 140/90, it was 13.0% (CI 5.6% - 19.8%).

**Conclusions:** The risk of incident hypertension after an HDP is high in the first year postpartum. Despite this, for many participants, BP was not even measured within 15-90 days postpartum despite. Opportunities exist to improve care for individuals with HDPs.

## Introduction

Cardiovascular disease (CVD) is the leading cause of death in women, and, though often preventable, CVD mortality among young women has disproportionately increased in recent years.^1^ Hypertensive disorders of pregnancy (HDP) include, in order of increasing severity, gestational hypertension without proteinuria, preeclampsia without severe features, chronic hypertension with superimposed preeclampsia, and severe preeclampsia/eclampsia. Each is a risk factor for long and short-term CVD disease including heart failure, myocardial infarction, stroke and CVD mortality.^2^ Administrative data from South Carolina births (2004-2016) shows that HDPs confer additional risk even within 5 years of delivery.^3^

The diagnosis and timely postpartum management of HDP provides an opportunity to improve the outcomes of future pregnancies and prevent long-term CVD morbidity and mortality. Pregnancy functions as a metabolic stress test, identifying a predisposition to CVD before it leads to morbidity and mortality.^4^ The diagnosis of HDPs provides a prevention opportunity for individuals who might otherwise not come to clinical attention early enough to prevent morbidity and mortality. Current guidelines for short- and long-term postpartum management of individuals with HDP are relatively recent and somewhat inconsistent.^5^ However, all organizations agree that management of CVD risk factors, such as hypertension, is important for individuals after an HDP. Currently, the American College of Obstetricians and Gynecologists recommends that individuals with HDPs have a blood pressure (BP) evaluation 3-10 days after delivery and that all patients should be “educated on their individual risk” and have a repeat BP evaluation at their comprehensive visit at 15-90 days postpartum with the goal of creating a postpartum management plan for hypertension.^6,7^ The American Heart Association recommends frequent cardiac risk factor screening assessments in the first year postpartum at 6 weeks, 12 weeks, 6 months, and 12 months, with appropriate transition from postpartum to longitudinal primary care around the 8- to 12-week mark.^8^

Identification and appropriate management of hypertension is one key aspect in the prevention of CVD and adverse pregnancy outcomes.^5^ Few studies investigate the risk of hypertension after an HDP in U.S. populations and those that have been conducted do not control for known risk factors for heart disease including race, ethnicity, smoking, and family history.^9^ It is also unclear whether different categories of HDP would confer a greater risk of incident hypertension and which other factors may modify this risk.^4^ Finally, very few examine the risk of incident hypertension in the first year after delivery. This makes it difficult for obstetrics and primary care providers to appropriately counsel patients after pregnancy “about their individual risk” or to employ specific preventive strategies to improve future pregnancy outcomes and long-term health of the individual.

In order to better understand the early risk of incident hypertension after an HDP event, the primary objective of this analysis was to estimate the associations between systolic and diastolic BPs during routine postpartum follow-up and the relative hazard of incident hypertension through the first year postpartum. We hypothesized that higher postpartum systolic and diastolic BPs would be associated with a higher incidence of incident hypertension diagnosis within 12 months. Secondary objectives were to estimate the associations between other potential risk factors for incident hypertension during the first year postpartum and to understand the proportion of individuals who had BP measurement per American College of Obstetricians and Gynecologists guidelines.

## Methods

### Study Design

This was a planned secondary analysis of a retrospective cohort study investigating factors associated with postpartum risk of hypertension among people 13-54 years old diagnosed with an HDP at two academic health systems. REporting of studies Conducted using Observational Routinely-collected health Data (RECORD) guidelines were followed. See **Supplement Table 1**.

### Source of Participants and Data

We used data from the Carolinas Collaborative, a partnership between the National Institutes of Health’s Clinical and Translational Science Award hubs in North and South Carolina which harmonizes the electronic health data across institutions to expedite clinical research.^10^ Using the common data model of the Carolinas Collaborative, we identified women with a hypertensive disorder of pregnancy and delivering at two academic health systems in the southeastern U.S. between January 1, 2007 and December 31, 2017. Each health system includes a main campus as well as smaller, community hospitals. Women 13-54 years old with an International Classification of Diseases, Ninth/Tenth Revision, Clinical Modification (ICD-CM 9/10) code for an HDP were included (ICD-10-CM O11, O13, O14, O15 and ICD-9 642.3, 642.4, 642.5, 642.6, 642.7). A team from each institution abstracted the data for eligible patients from their respective electronic health records. They then shared the data with each other to match patients who had records in both health systems using a hashing algorithm which contained the first three digits of the first name, first eight digits of the last name, gender, and date of birth. The data was merged and de-identified except dates of service, dates of birth, and zip codes.

For the purposes of this analysis, we included HDP-affected deliveries which occurred only in 2014 or later and had a BP measured 15-90 days postpartum in either system by any provider. We excluded data before 2014 in this analysis due to a high proportion of missing data, especially BP measurements. The Epic, Epic Systems Corporation rolled out in both institutions in 2014 and likely contributed to higher data quality after that point. In women who had multiple recorded pregnancies complicated by an HDP, only the first pregnancy in the time period was included. We excluded records that had improbable gestational age at delivery (<0 or >43 weeks). Given our focus on incident hypertension related to postpartum BP measurement, we excluded patients who had a diagnosis of pre-existing hypertension before or during pregnancy or at the time of the first BP measurement during the 15-90 day postpartum window. Per national and international guidelines, the diagnosis of incident hypertension cannot be accurately made before 12 weeks of gestation due to transient changes in BP that occur during this period.^11–14^

### Variables of Interest

The primary outcome was time to diagnosis of hypertension by any provider (ICD-10-CM I10 and I11; ICD 9 401 and 402) after the date of the 15-90 day BP check. Participants were censored at end of study on December 31^th^, 2019. The primary exposure was systolic and diastolic BP at the comprehensive postpartum visit ^15^ We included several potential predictors of time to incident hypertension diagnosis, including type of HDP, gestational age of HDP onset, year of delivery (2014 – 2017), days from delivery to BP measurement, maternal age at delivery, race (White, Black, or Other) and ethnicity (Hispanic or non-Hispanic), payor (commercial, government, or other [including self-pay]), median household income of ZIP code of maternal residence, marital status (divorced, married, single, or other), maternal body mass index (BMI), parity, presence of gestational diabetes mellitus, history of Type 1, Type 2, or prediabetes diabetes (ICD-10-CM E10, E11 or ICD-9 250) prior to delivery, and smoking status prior to delivery (current, former or never). Chronic kidney disease was rare in our population and was not included. Family history of heart disease was not available in the data set. Race and ethnicity were captured from the demographic information in the electronic health record. Although ideally those capture the patient-identified race/ethnicity, it is unclear to what extent race and ethnicity may have been misassigned within the electronic health record.^16^ When defining type of HDP, if women had multiple HDPs diagnosed during the pregnancy, we used the most severe diagnosis available. Hierarchical severity was: 1) gestational hypertension (ICD-10-CM O13*; ICD-9-CM 642.3*) 2) preeclampsia (ICD-10-CM O14.0* and O14.9*; ICD-9-CM 642.4*), and 3) eclampsia/severe preeclampsia (ICD-10-CM O14.1* and O14.2*, O15*; ICD-9-CM 642.5* and 642.6*). We combined the categories of eclampsia and severe preeclampsia because the clinical management of these diagnoses is similar, and the group sizes were small. To determine the time of onset of HDP, we used the time to the onset of the first HDP, not necessarily the most severe.

### Data Analytic Approach

Summary statistics of variable distributions were calculated. We used Cox proportional hazards models to estimate hazard ratios (HRs) for diagnosis of incident hypertension associated with systolic and diastolic BP at 15-90 days postpartum. The proportional hazards assumption was checked using graphical methods. Approximately 31% of the observations had missing values for at least one covariate. Therefore, we created 31 multiple imputed datasets and combined the estimates from each model according to Rubin’s Rules. From the fitted Cox model, we estimated a cumulative incidence of hypertension diagnosis postpartum, with otherwise typical covariate values for this cohort (median for continuous; most common for categorical). Because many BP measurements had a missing provider, we imputed the provider type using single imputation. For example, if an encounter was missing the provider type, we randomly assigned the encounter to be included in or excluded from the analysis set based on a Bernoulli random variable with probability equal to the sample proportion of encounters that did have the target provider type (i.e., primary care or obstetrics) assuming the proportion of encounters with the target provider type was the same for missing and non-missing providers. Models were fit using the R statistical computing environment,^17^ including the rms package.^18^ Multiple imputation was performed using the mice package.^19^

We conducted sensitivity analyses based on the provider specialty when the BP check was performed. In the first sensitivity analysis, we included only patients who had a BP measurement 15-90 days postpartum by a provider with the specialty of Obstetrics and Gynecology, Certified Nurse Midwife, Maternal and Fetal Medicine, Family Medicine, Obstetrics, Internal Medicine (including all subspecialties), or Pediatrics. In the second sensitivity analysis, we included only those with BPs taken by obstetric caregivers.

Programming code can be made available via request to the corresponding author.

### IRB Statement

This study was approved by the institutional review boards at both health systems, as well as by Auburn University where the data analysis was conducted. No informed consent was required.

## Results

There were 9,230 individuals in the original cohort. Of those, 5607 delivered between 2014-2017. After applying the remaining exclusion criteria, the final sample size for this analysis was 2,514. See **Figure 1**. Almost 40% (2125) of those individuals diagnosed with an HDP between 2014 and 2017 did not have a documented BP check by any clinician at 15-90 days postpartum in either of the healthcare systems. Of the women who would have otherwise met the inclusion criteria for the study, 146 were excluded because they had a diagnosis of hypertension before or on the day of the 15-90-day BP measurement. For details about characteristics of included versus excluded participants, see **Supplement Table 2**. Included participants were slightly older, less likely be Hispanic, but more likely to be White, married, have gestational diabetes, live in higher income ZIP codes, and have private insurance. Of the included participants, only 20% had a BP check at 7-14 days postpartum.

**Figure 1.**
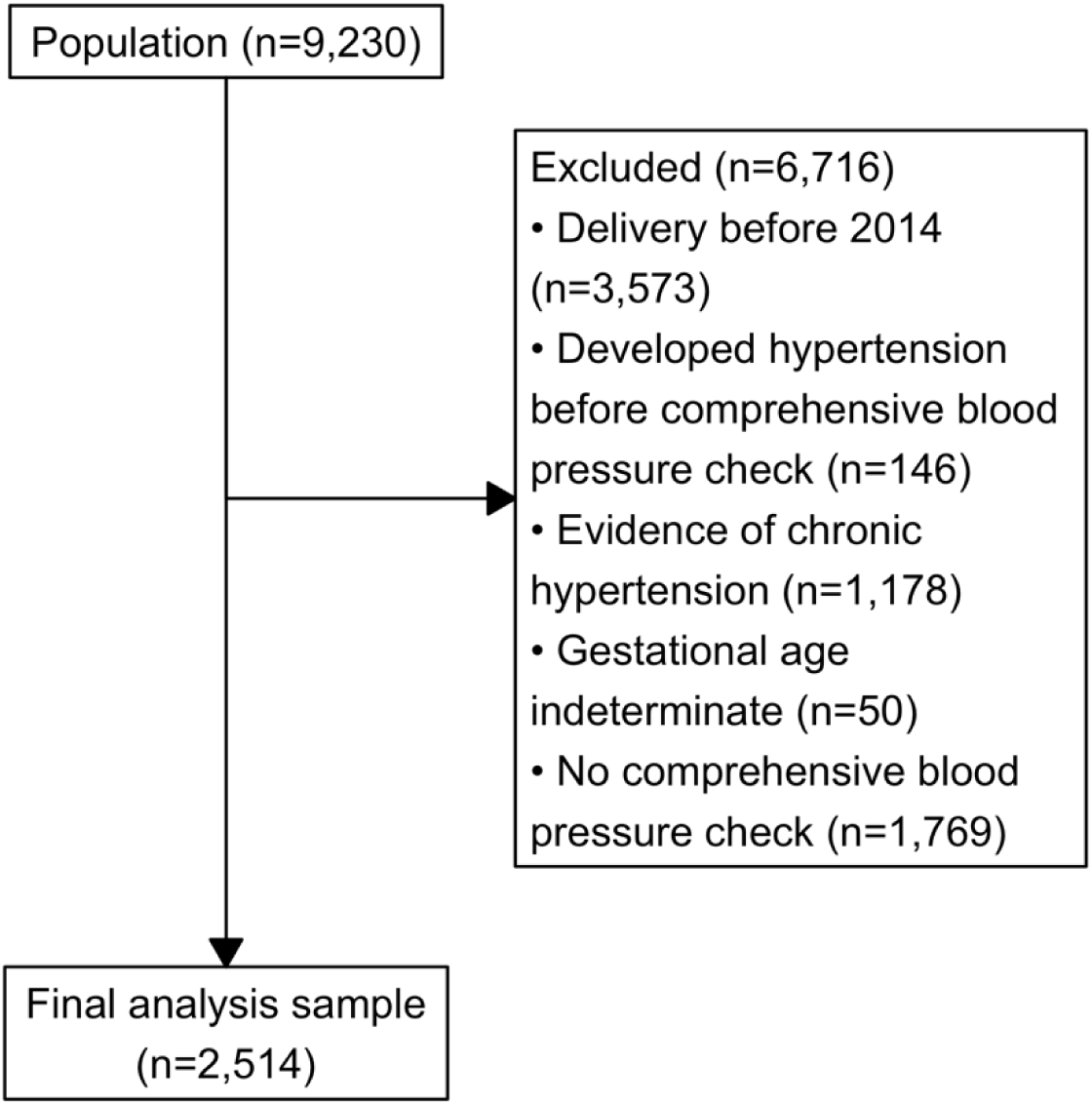
Study Flow Diagram of Included Patients for Analysis of Incident Hypertension After an HDP.

Summary statistics for each variable of interest according to American Heart Association BP ranges at 15-90 days postpartum are reported in **Table I**.^15^ Only 32% (799/2514) had BPs in the “normal” range and the majority (55%) had BPs in the range that would be classified as of Stage 1 or 2 Hypertension if similar or higher readings were obtained on a 2^nd^ occasion. Twenty percent also had a clinical BP measurement recorded within the first 14 days postpartum. Among those who did, almost 80% had BPs in the Stage 1 or 2 hypertension range. Thirteen percent of the population (315 participants) was diagnosed with incident hypertension after the 15-90-day BP measurement.

**Table I.**
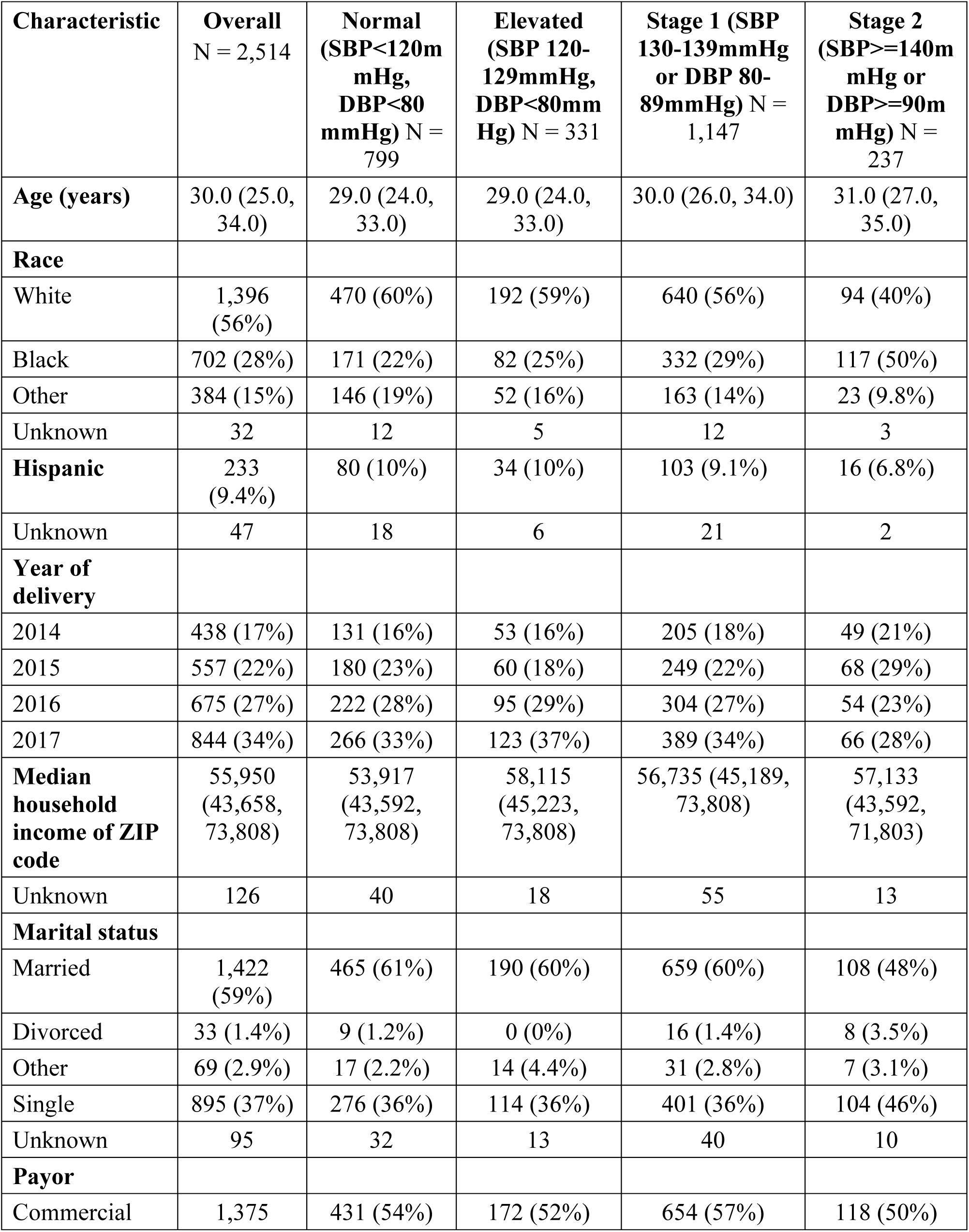
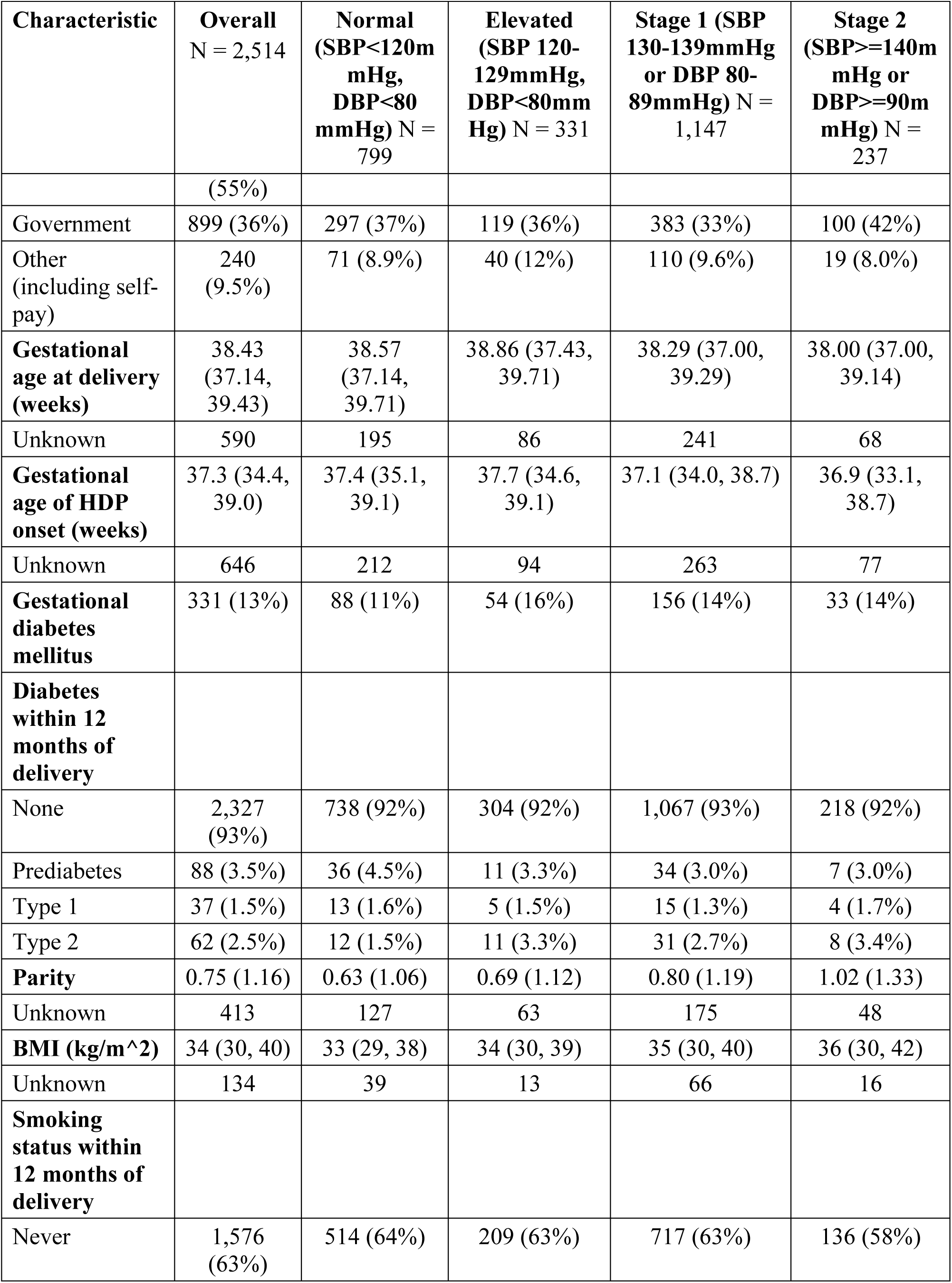
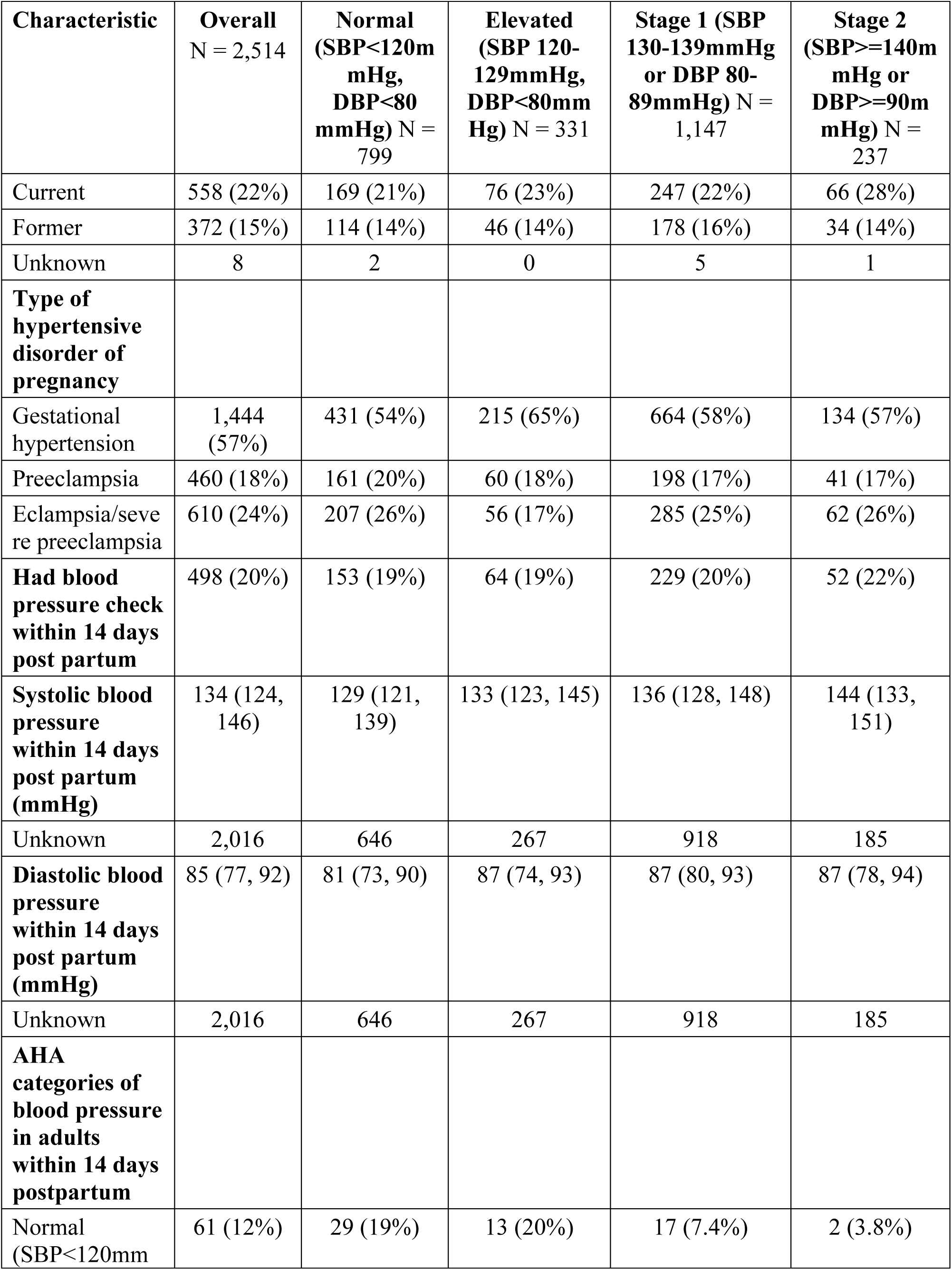
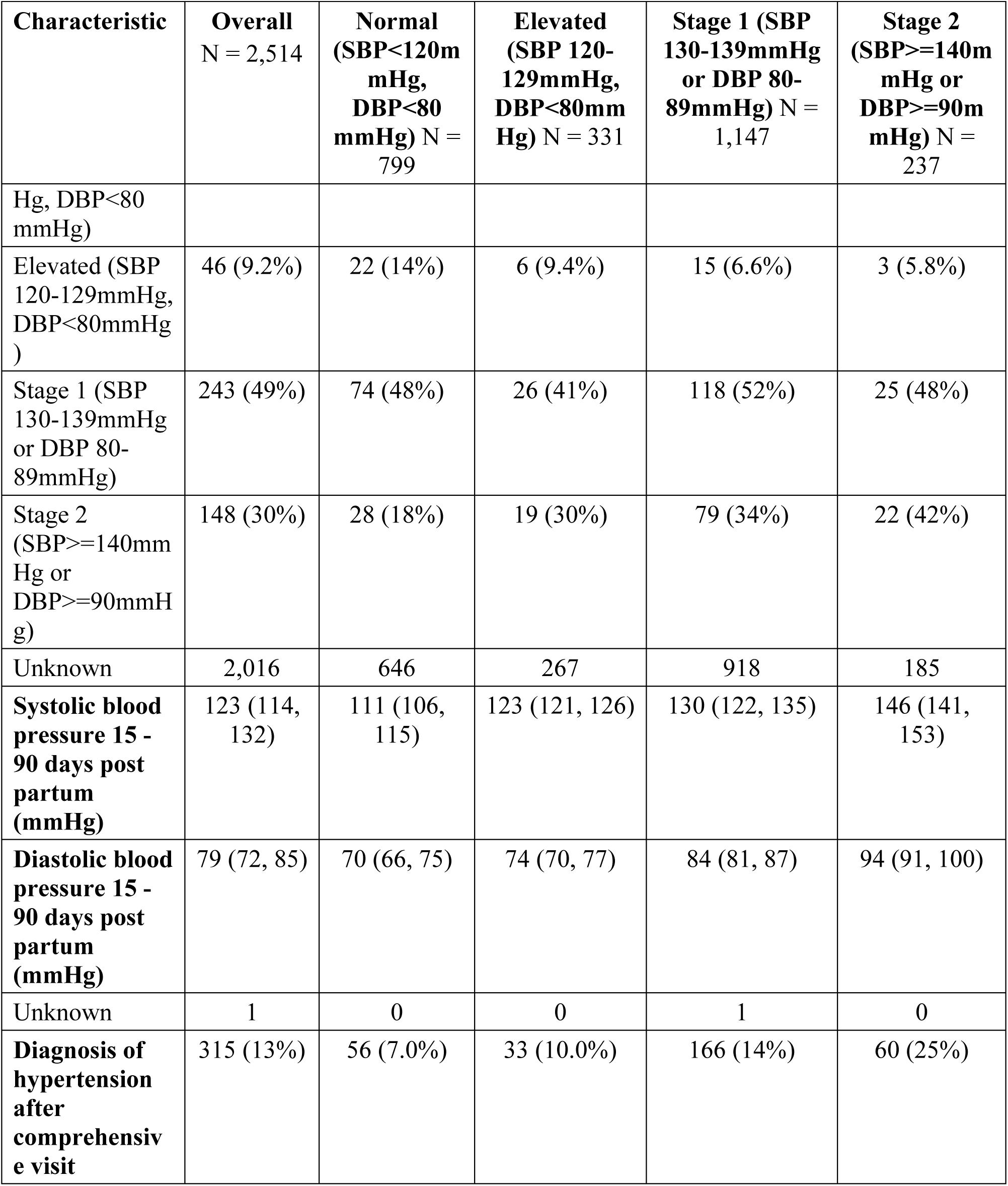
Description of women diagnosed with an HDP between 2014 and 2017 by American Heart Association BP categories 15-90 days postpartum. ^15^

A forest plot of the estimated HRs for incident hypertension for each Cox model covariate is provided in **Figure 2**. The HR of developing incident hypertension was statistically significantly higher for those with elevated (140 v 120 mmHg) systolic BP (HR 1.70 95% CI 1.37 - 2.12), for those with severe preeclampsia versus gestational hypertension (HR 1.49 CI 1.15 – 1.94), and older maternal age (35 versus 25 years, HR 1.46 CI 1.16-1.83). A longer time until the postpartum BP check (90 days v. 42 days) was associated with a lower hazard of incident hypertension (HR 0.52 CI 0.34-0.81). See **Table 2** for more details. No covariates demonstrated strong deviations from the proportional hazards assumption (see **Supplement Figure 2**). In the sensitivity analyses that restricted to only women whose comprehensive visit BP was performed by: (1) an obstetric or primary care specialty provider; or (2) an obstetric provider, the results were similar. See **Supplement Table 3** and **Supplement Figure 1**.

**Figure 2.**
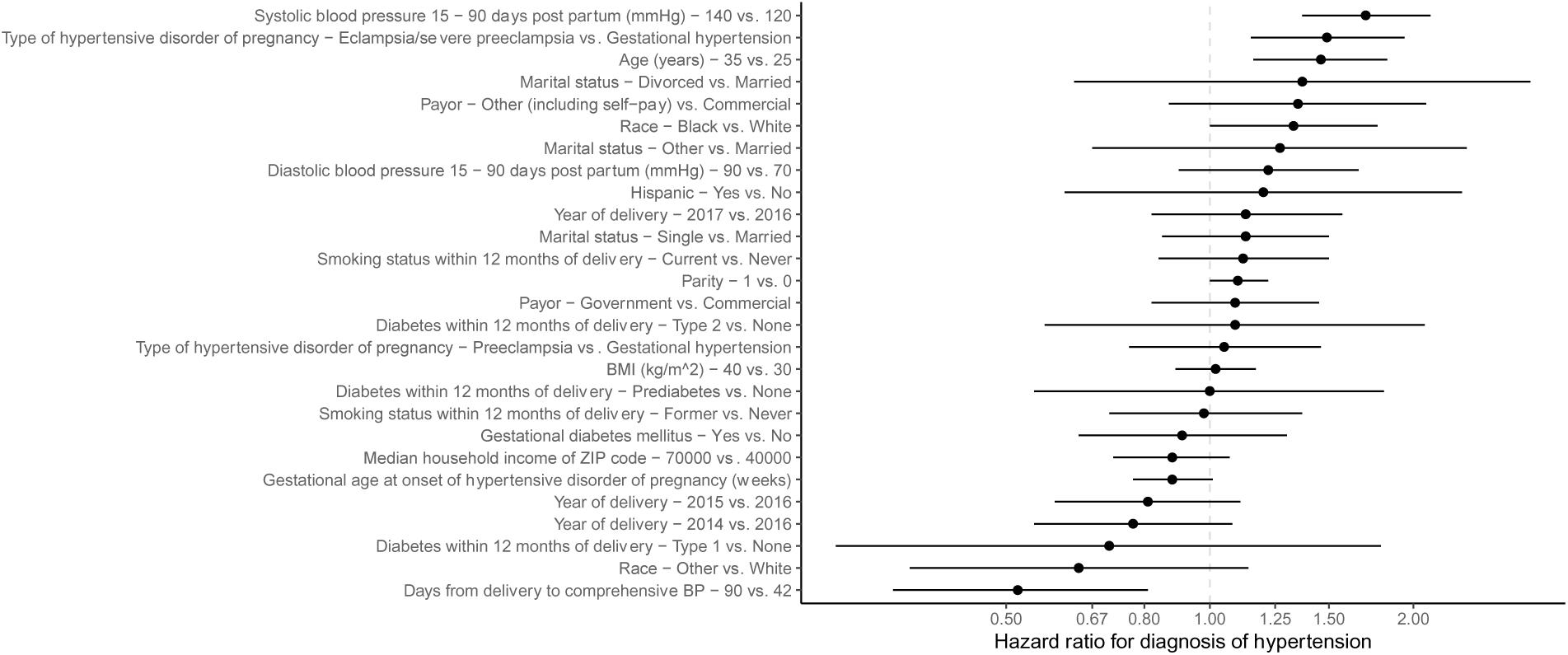
Hazard ratios and 95% confidence intervals for diagnosis of incident hypertension among women diagnosed with an HDP (n = 2514, m = 315 events).

**Table 2.** Incident hypertension diagnosis hazards among women diagnosed with an HDP at two academic medical centers (n = 2514, m = 315 events).

| Covariate | HR (95% confidence interval) |
| --- | --- |
| Systolic blood pressure 15 - 90 days post partum (mmHg) - 140 vs. 120 | 1.7 (1.37 - 2.12) |
| Diastolic blood pressure 15 - 90 days post partum (mmHg) - 90 vs. 70 | 1.22 (0.9 - 1.66) |
| Days from delivery to comprehensive BP - 90 vs. 42 | 0.52 (0.34 - 0.81) |
| Age (years) - 35 vs. 25 | 1.46 (1.16 - 1.83) |
| Median household income of ZIP code - 70000 vs. 40000 | 0.88 (0.72 - 1.07) |
| BMI (kg/m<sup>2</sup>) - 40 vs. 30 | 1.02 (0.89 - 1.17) |
| Parity - 1 vs. 0 | 1.1 (1 - 1.22) |
| Gestational age at onset of hypertensive disorder of pregnancy (weeks) | 0.88 (0.77 - 1.01) |
| Type of hypertensive disorder of pregnancy - Preeclampsia vs. Gestational hypertension | 1.05 (0.76 - 1.46) |
| Type of hypertensive disorder of pregnancy - Eclampsia/severe preeclampsia vs. Gestational hypertension | 1.49 (1.15 - 1.94) |
| Year of delivery - 2014 vs. 2016 | 0.77 (0.55 - 1.08) |
| Year of delivery - 2015 vs. 2016 | 0.81 (0.59 - 1.11) |
| Year of delivery - 2017 vs. 2016 | 1.13 (0.82 - 1.57) |
| Race - Black vs. White | 1.33 (1 - 1.77) |
| Race - Other vs. White | 0.64 (0.36 - 1.14) |
| Hispanic - Yes vs. No | 1.2 (0.61 - 2.36) |
| Payor - Government vs. Commercial | 1.09 (0.82 - 1.45) |
| Payor - Other (including self-pay) vs. Commercial | 1.35 (0.87 - 2.09) |
| Marital status - Divorced vs. Married | 1.37 (0.63 - 2.98) |
| Marital status - Other vs. Married | 1.27 (0.67 - 2.4) |
| Marital status - Single vs. Married | 1.13 (0.85 - 1.5) |
| Gestational diabetes mellitus - Yes vs. No | 0.91 (0.64 - 1.3) |
| Smoking status within 12 months of delivery - Current vs. Never | 1.12 (0.84 - 1.5) |
| Smoking status within 12 months of delivery - Former vs. Never | 0.98 (0.71 - 1.37) |
| Diabetes within 12 months of delivery - Prediabetes vs. None | 1 (0.55 - 1.81) |
| Diabetes within 12 months of delivery - Type 1 vs. None | 0.71 (0.28 - 1.79) |
| Diabetes within 12 months of delivery - Type 2 vs. None | 1.09 (0.57 - 2.08) |

**Figure 3** shows the modeled cumulative incidence of hypertension within 12 months after the 15-90 day BP check for specific postpartum BPs categories: optimal, at-risk, Stage 1 hypertension and Stage 2 hypertension.^15,20^ The other covariates in the model were set at the typical values in our sample (i.e., median values for continuous and most common values for categorical variables). For women with a BP in the optimal range (110/65 mmHg), the risk of being diagnosed with hypertension by 12 months after the 15-90 day BP measurement was 4.7% (CI, 2.0% - 7.4%). For women with a systolic BP of 140 mmHg and diastolic BP of 90 mmHg, the risk was 13.0% (CI 5.6% - 19.8%).

**Figure 3.**
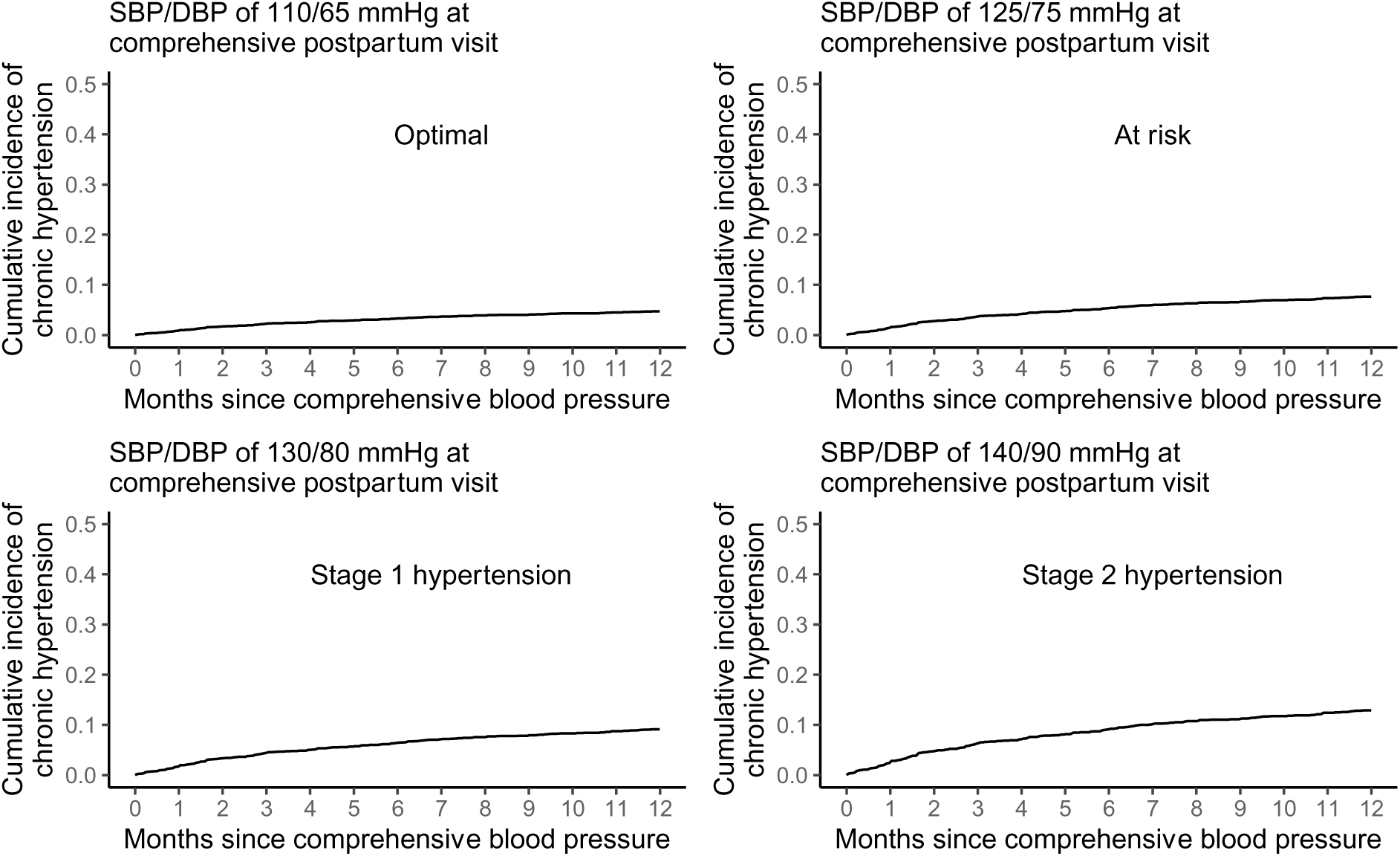
Hazard ratios and 95% confidence intervals for diagnosis of incident hypertension after an HDP(n = 2514, m = 315 events), by postpartum BP category. Other covariates were set to typical values in the dataset, so these predicted cumulative incidences are for a hypothetical woman with a non-severe clinical profile. For example, these predictions are for a non-Hispanic White nulliparous woman age 29, with a diagnosis of preeclampsia at 37 weeks, with no history of diabetes or gestational diabetes, a never smoker, and a BMI of approximately 35 kg/m2.

## Discussion

Among women diagnosed with HDPs at two academic health systems, more than 1 in 10 women developed incident hypertension by 12 months after delivery. Higher systolic BP at the postpartum visit, older age, and more severe HDP were associated with a greater risk. The proportion of individuals without a 15-90 day postpartum BP was high (40%) despite our inclusion of any BP completed during any clinical interaction in the two health systems. The majority of the 15-90 day postpartum BP evaluations were conducted by obstetrical providers. During the study period, the early BP screening recommendation for individuals who experience HDPs was relatively new, the guideline having been first published in November 2013.^21^ Our findings are consistent with other studies in that only about 40-60% of patients with HDPs have a postpartum BP check during the first 90 days postpartum.^22–24^

There is relatively little data in U.S. populations about the risk of developing hypertension in the year after a delivery complicated by an HDP. In a Danish cohort (1995-2007), the hazards of developing hypertension in the year after delivery was 12 to 25-fold higher than for individuals without an HDP. The hazards ratios were lower for later time points, suggesting that the first year after pregnancy may be a particularly common time to diagnose hypertension.^25^ As in our study, adjusting for pre-existing diabetes did not have an impact on the outcome and increasing severity of the HDP was associated with an increased risk of incident hypertension.^25^ In a Scottish study (1951-1970) increasing severity of the HDP was associated with higher odds of incident hypertension was (1.95 for gestational hypertension and 2.97 for preeclampsia/eclampsia).^26^ We found no differences in the hazards of incident hypertension by race, income, and insurance status, and there are very little data investigating these associations.^27,28^ Our group has also demonstrated that early BP screening is associated with increased detection of other incident cardiovascular risk factors.^29^

The strengths of our study include a large sample size allowing us to follow patients with HDPs for up to 4 years after delivery. The study used routinely collected electronic health data and thus is reflective of real-world clinical practice and applicable to others seeking to use such data to perform population surveillance and quality improvement. Our population was diverse by race, ethnicity, zip code median income, clinical risk factors for hypertension and insurance status, which is rare in studies of HDPs.^28^

This study was limited by the retrospective nature of the data analysis. Additionally, patients who delivered in these health systems may have been transferred from geographically distant areas in the state specifically because of their diagnosis of HDP and may have subsequently received postpartum care back in their own community leading to missing BP measurements for those individuals. A third limitation in the study is that individuals included in the study were older, more likely to be White, less likely to be Hispanic, more likely to be married, more likely to be commercially insured, and less likely to smoke than individuals excluded from the study. Many of these differences are associated with better access to health care in general and likely impacted these individuals’ ability to receive recommended postpartum care.^30,31^ However, around half of exclusions were due to preexisting hypertension which likely partially explains the differential distribution of CVD risk factors (i.e. age and smoking). A fourth limitation is that our outcome of incident hypertension was a clinical diagnosis that relied upon ICD-10-CM codes. Use of these codes to identify hypertension is consistent with expected population estimates of hypertension and is frequently used as an outcome in studies of long-term risk after HDP.^32,33^ The potential for misclassification of HDPs by ICD-10-CM codes also exists; however, validation studies have shown that these codes used together have very high (>95%) specificity and sensitivity for any HDP and very high specificity for each subcategory.^34^ The hazards of receiving an ICD-10-CM diagnosis of hypertension may be impacted by clinical decision-making. For example, older women may have been more likely to be given the diagnosis of hypertension by their providers than younger women with the same BP measurements. The finding that different race and ethnicity categories were not associated with differential risk of incident hypertension is limited in that these individuals were less like to obtain a postpartum BP check, and there are well-documented disparities in accessing postpartum BP checks for non-Hispanic Black and Hispanic patients.^35,36,24^ This study may not be generalizable not in the U.S. South.

Our study has several important implications for future policy related to appropriate postpartum care. First, current clinical guidelines for postpartum BP evaluation in individuals with HDP were not consistently applied. There are health system level barriers (e.g. insufficient patient education, difficulty accessing appointments) and patient-level barriers (e.g. transportation, caring for newborn) to receiving this care.^31,37^ Opportunities exist for use of electronic health data to identify eligible patients and offer BP screening after an HDP when it has not yet occurred. This could be paired with programs to remove barriers for patients such as home monitoring, telehealth, and/or home visitors.^38,22^ Second, given the significant numbers of individuals diagnosed with incident hypertension regardless of severity of HDP or BP measurement postpartum, preventive guidelines for long-term health should be consistent and should prioritize early lifestyle modification and control of BP and other CVD risk factors. All providers who see patients who have experienced HDPs should be aware of potential risks and should identify and manage hypertension early. Future research should explore preventive interventions to improve hypertension management for individuals who experience HDPs, ideally codesigned with patients so as to address the unique needs and experiences of the postpartum period.^39,40^

## Data Availability

Data can be made available upon requests to the authors.

## Non-Standard Abbreviations and Acronyms

CVD: Cardiovascular Disease
HDP: Hypertensive Disorder of Pregnancy
BP: Blood Pressure
CI: 95% Confidence Interval
ICD-CM 9/10: International Classification of Diseases, Ninth/Tenth Revision, Clinical Modification
BMI: Body Mass Index
HR: Hazard Ratio

## Acknowledgments

We want to acknowledge Amanda Stebbins from the Duke Clinical Research Assistant for her help with data management.

